# Assessing Benefit in Heart Failure Patients with Reduced Ejection Fraction: Analysis of the VICTORIA Trial Using Novel Prognostic Risk Stratification

**DOI:** 10.1101/2025.04.03.25325054

**Authors:** Christopher M. O’Connor, Sarah Rathwell, Devan V. Mehrotra, Stefano Corda, Ciaran J. McMullan, Carolyn S.P. Lam, Justin A. Ezekowitz, Burkert Piekse, Adrian F. Hernandez, Kevin J. Anstrom, Robert J. Mentz, Christopher R. deFilippi, Adriaan Voors, Piotr Ponikowski, Javed Butler, Cynthia M. Westerhout, Paul W. Armstrong, the VICTORIA Study Group

## Abstract

**Background:** Randomized controlled clinical trials remain the gold standard for determining efficacy of new heart failure (HF) therapies; however, failure to account for heterogeneity in risk of the primary endpoint(s) may dilute treatment efficacy. The novel 5-step stratified testing and amalgamation routine (5-STAR) methodology addresses these limitations using risk stratification based on treatment-independent associations between baseline covariates and clinical outcomes. We applied the 5-STAR methodology to the original VICTORIA database enriched by relevant ancillary information.

**Methods:** Within the 5-STAR analysis, elastic net Cox regression and a conditional inference tree tool blinded to treatment assignment were used to partition the trial population into risk strata for trial endpoints based on baseline covariates determined to be jointly strongly associated with the risk of the outcome. Core laboratory mechanistic biomarkers and baseline electrocardiographic variables were added to the VICTORIA dataset. After unblinding, treatments were compared for the primary composite endpoint of cardiovascular death or HF hospitalization within each risk stratum: stratum-level results were then averaged for overall inference.

**Results:** The 5-STAR analysis showed a greater vericiguat treatment effect on the primary composite endpoint than the original prespecified VICTORIA analysis (5-STAR-averaged HR, 95% CI: 0.85, 0.77–0.94 vs 0.90, 0.82–0.98), and on its components (5-STAR-averaged HR, 95% CI: cardiovascular death: 0.79, 0.67–0.93 vs 0.93, 0.81–1.06; HF hospitalization: 0.89, 0.79–1.00 vs 0.90, 0.81–1.00). Five biomarkers (GDF-15, NT-proBNP, albumin, blood urea nitrogen, urate) determined the risk strata across the 3 endpoints.

**Conclusions:** By developing treatment-independent risk stratification, the 5-STAR methodology attenuates dilution of treatment effects inherent in conventional prognostic risk heterogeneity. This retrospective analysis of VICTORIA revealed greater efficacy of vericiguat on the primary endpoint and its components. GDF-15 was consistently the strongest prognostic risk factor across the composite endpoint and its components of cardiovascular death and HF hospitalization.

**Clinical Trial Registration:** ClinicalTrials.gov (NCT02861534).

## INTRODUCTION

Despite recent therapeutic advances in patients with high-risk heart failure and reduced ejection fraction (HFrEF), morbidity and mortality remain high.(1–3) In the VICTORIA trial, vericiguat, a soluble guanylate cyclase stimulator, compared with placebo reduced the composite endpoint of cardiovascular death or HF hospitalization in patients with HFrEF recently hospitalized with HF.(2) In additional efficacy analyses across 13 pre-specified, one-factor-at-a-time subgroups, a beneficial effect of vericiguat was observed in the lower 3 quartiles of baseline N-terminal pro-B-type natriuretic peptide (NT-proBNP) levels but not in the upper fourth quartile.

Randomized controlled clinical trials remain the gold standard for determining efficacy of new HF therapies; however, risk heterogeneity-especially when discovered after the trial is completed-may dilute treatment efficacy thereby obscuring which patients benefit from therapeutic interventions. The original efficacy analyses for VICTORIA used a stratified Cox model based on two pre-selected stratification factors: region and race. However, analyses based on suboptimal stratification such as these (i.e., based on factors/covariates that are weakly prognostic for clinical outcomes) can dilute treatment efficacy signals.(4) The 5-step stratified testing and amalgamation routine (5-STAR) methodology provides a novel safeguard against this potential dilution. Using the observed data for each clinical outcome of interest, a treatment-blinded algorithm partitions the heterogenous patient population into well-separated risk strata defined by baseline covariates that are determined to provide a strongly prognostic template for outcome risk. After this process is completed, treatment unblinding occurs and comparison is done within each risk stratum. Stratum-level results are then averaged for overall inference.(5) Improved risk stratification achieved with 5-STAR thereby potentially boosts power for detecting treatment efficacy.

In the current analysis, we applied the 5-STAR methodology to the VICTORIA trial including an expanded pre-specified set of candidate risk variables such as mechanistic biomarkers and electrocardiographic measures assessed at core laboratories. Our intent was to gain additional insight into the risk heterogeneity of the population we studied and apply those insights to vericiguat’s treatment effect on the primary composite outcome of cardiovascular death or HF hospitalization and each of the components. A comparison of results using 5-STAR (based on identified prognostic factors that defined risk strata) versus the originally reported analysis (based on pre-specified stratification factors) was also conducted.

## METHODS

### Patient Population

The design and results of the VICTORIA trial have been published.(2,6) In brief, VICTORIA was a randomized trial comparing vericiguat with placebo in 5050 patients with chronic HFrEF. The primary endpoint was the composite of cardiovascular death or HF hospitalization; secondary endpoints included the components of the primary endpoint.

This study is a retrospective analysis from the VICTORIA trial published in the *New England Journal of Medicine*.(2) The trial protocol was approved by regulatory agencies in the participating countries and institutional review boards or ethics committees at the participating sites.

### 5-STAR Methodology

Patient characteristics, physical exam measurements, laboratory values, and health status were collected using standardized forms at baseline. Specific instructions and definitions for all variables were provided to assist the sites with form completion. Mechanistic quantitative biomarkers and electrocardiographic variables were measured in validated core laboratories.(7,8)

The 5-STAR methodology applies 5 consecutively ordered steps; removal of high missingness variables (step 1), removal of noise variables (step 2), partitioning of treatment-agnostic risk strata (step 3), treatment effect estimation within risk strata (step 4), and total treatment effect pooling (step 5). In total, 82 baseline variables were candidates for potential association with each clinical endpoint of interest (**Table S1**) in step 1 of 5-STAR.They included the 66 variables considered in risk stratification models previously developed in the placebo group(9) and were selected as most meaningful and formed the basis for the main covariates. In addition, 5 mechanism-specific biomarkers(7) and 11 electrocardiographic variables(8) were included, bringing the total of pre-specified candidate variables identified for possible risk stratification to 82. No transformations of the variables were performed, and missing data were not imputed as the 5-STAR algorithm handles missingness internally. Variables with <10% missingness, or 10–20% missingness with an association to the outcome variable (log-rank p-value <0.05) were passed and progressed forward through the initial filtering, pre-processing step. No variables were removed from the models due to high missingness. Three variables had >10% missingness but were sufficiently associated with all endpoints (high-sensitivity C-reactive protein: 10.5%, cystatin C: 10.8%, growth differentiation factor-15: 13%) and were included.

“Noise” variables (i.e., those without sufficient relationship to the endpoint of interest, blinded to treatment arm) were then eliminated by elastic net Cox regression to provide a subset of variables significantly associated with the outcome (step 2 of 5-STAR). Because these models were not developed for predictive or prognostic purposes discriminatory power or calibration were not assessed. A conditional inference tree algorithm then used the shortened variables list subsequently used by a conditional inference tree tool (C-tree) and an unbiased recursive-partitioning program(10) was used to segregate the overall risk-heterogenous patient population into subpopulations composed of homogeneous patients relative to risk strata (step 3). A preliminary tree was constructed to categorize participants into a maximum of 4 risk groups with a minimum group size of 5% of the total sample size as previously applied and based on the variables passed through the previous step (4). For variables with missing data, up to 3 surrogate variables were analyzed to determine how to classify participants. A final tree was then constructed by combining preliminary risk strata that were not sufficiently distinct (p-value ≥0.2). Within each risk stratum, a treatment comparison of vericiguat versus placebo was conducted using Cox regression for each of the 3 endpoints (step 4). An overall statistical inference was obtained by pooling the stratum-level findings, accounting for the relative size of each risk stratum and variability of the estimate (step 5), as previously detailed.(5) The primary focus was on the estimated overall (i.e., risk-stratum-averaged) hazard ratio (HR) and corresponding 95% confidence interval (CI). The probability of treatment benefit (Prob HR<1) was also calculated, overall and for each risk stratum, and the average HR using 5-STAR was contrasted with the HR reported in the original prespecified VICTORIA analysis. The analyses were performed with the R package fiveSTAR, version 0.1.0.(4,5)

## RESULTS

Between 2016 and 2018, 5050 patients were enrolled in the VICTORIA trial at 616 centers across the globe. There were no significant differences in baseline patient characteristics between treatment and placebo arms, and the median follow-up was 10.8 months. There was a total of 1869 primary endpoints comprising 431 cardiovascular deaths and 1438 HF hospitalizations as first events; 855 patients experienced the secondary endpoint of cardiovascular death.(2) The original prespecified analysis revealed a 10% relative reduction (HR 0.90, 95% CI 0.82–0.98; p=0.02) in the primary endpoint. There was a statistically significant interaction with quartiles of plasma NT-proBNP suggesting lack of benefit of vericiguat in the uppermost quartile (HR 1.16, 95% CI 0.99–1.35).(2) A treatment benefit with vericiguat was also observed for HF hospitalization (HR 0.90, 95% CI 0.81–1.00) but not CV death (HR 0.93, 95% CI 0.81–1.06).

### Risk Strata Development

From the list of 82 candidate variables, GDF-15 (according to a cut point of 4265 pg/mL) and subsequently NT-proBNP emerged as the most important variables for stratifying risk of the composite endpoint. For the assessment of cardiovascular death GDF-15 was also the most important risk-determining variable followed by NT-proBNP and serum albumin in subsequently defining risk. The final risk strata tree for HF hospitalization was also first defined by GDF-15 followed by serum uric acid and blood urea nitrogen (BUN) which constituted the second most important variables for defining HFH risk. All 4 preliminary risk strata for each endpoint were passed through to the second tree to determine if any groups were similar enough to be pooled. Both the composite and cardiovascular death had two preliminary strata that were homogenous enough to be combined (p-values ≥0.2) yielding 3 final risk strata each for these endpoints (**Figures S1 and S2**). All preliminary risk strata for heart failure hospitalization all showed enough heterogeneity to be passed through the final tree yielding 4 final risk strata (**Figure S3**).

### Clinical Outcomes Across Risk Strata and Vericiguat’s Treatment Effect

In **Figure 1** (left panel), the Kaplan Meir curves for the 3 risk strata identified for the primary composite endpoint are shown. These reveal a wide spectrum of event-free probabilities within the observed time period of the trial ranging from 63% likelihood of no event in the lowest risk group to 25% in the highest risk group. In the right panel, a comparison of event rates for the vericiguat and placebo groups is shown. Vericiguat demonstrated an overall 5-STAR-averaged 15% reduction in the primary endpoint versus placebo (HR 0.85, 95% CI 0.77–0.94). In **Figure 2** (left panel), there was also a wide spectrum of event-free probabilities of cardiovascular death occurrence in the 3 strata that ranged from 84% to 48%. In **Figure 2** (right panel), vericiguat was associated with a 21% reduction in the risk of cardiovascular death as compared with placebo (5-STAR-averaged HR 0.79, 95% CI 0.67–0.93). The range of event-free probabilities in the 4 risk strata of HF hospitalization ranged from 73% to 38% likelihood of no event (**Figure 3** [left panel]). Vericiguat demonstrated an 11% reduction in HF hospitalization (5-STAR-averaged HR 0.89, 95% CI 0.79–1.00) (**Figure 3** [right panel]).

**Figure 1.**
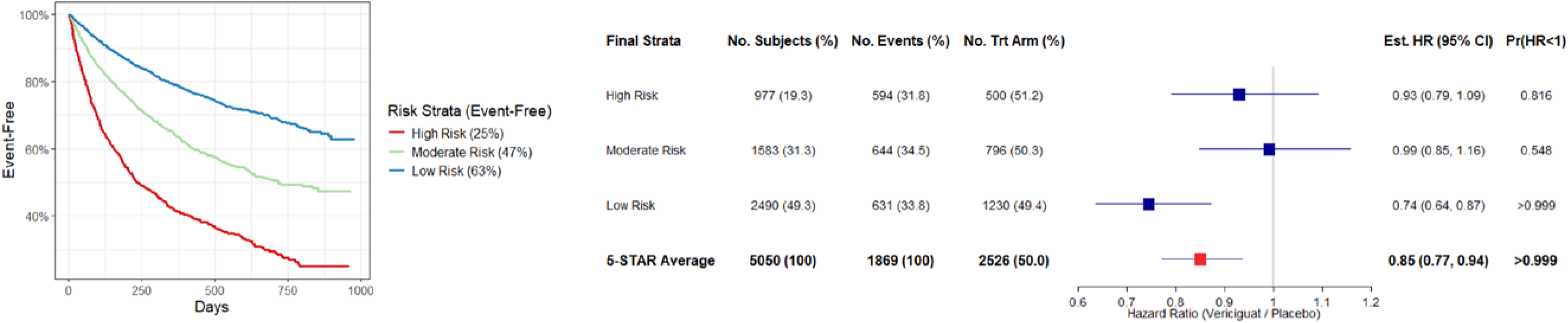
Risk Strata for the Primary Composite of Cardiovascular Death (CVD) and Heart Failure-hospitalization (HFH) (right), and Strata-specific and 5-STAR Averaged Treatment Effect (vericiguat versus placebo) (left). CI indicates confidence interval; HR, hazard ratio; Pr(HR<1), probability of vericiguat’s benefit.

**Figure 2.**
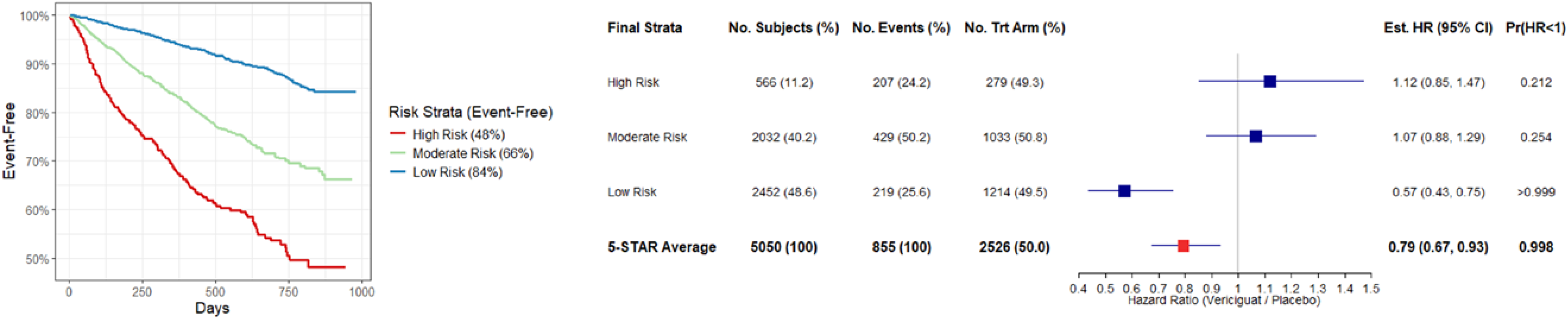
Risk Strata for Cardiovascular Death (CVD) (right), and Strata-specific and 5-STAR Averaged Treatment Effect (vericiguat versus placebo) (left). CI indicates confidence interval; HR, hazard ratio; Pr(HR<1), probability of vericiguat’s benefit.

**Figure 3.**
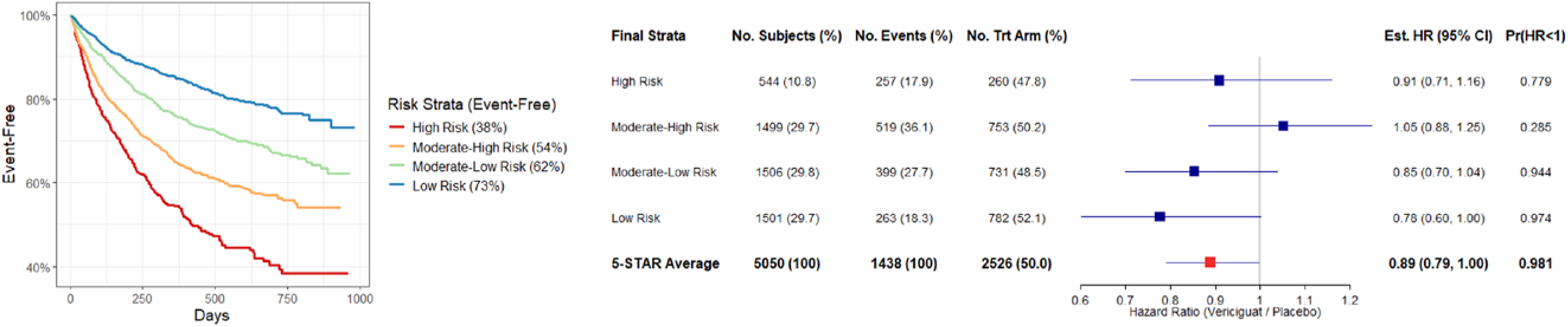
Risk Strata for Heart Failure-hospitalization (HFH) (right), and Strata-specific and 5-STAR Averaged Treatment Effect (vericiguat versus placebo) (left). CI indicates confidence interval; HR, hazard ratio; Pr(HR<1), probability of vericiguat’s benefit.

These observations derived from the 5-STAR methodology are in contrast to the original aforementioned findings of vericiguat’s effect on the hazard ratios for the composite endpoint and for cardiovascular death and to a lesser degree for HF hospitalization.

## DISCUSSION

There are three major findings from this analysis. First, and most importantly, the 5-STAR methodology provides an analytical approach that yields better precision on the efficacy of an intervention based on the alignment between baseline characteristics and outcomes. In the case of VICTORIA this approach demonstrated a stronger efficacy signal favoring vericiguat over placebo, for the primary composite endpoint of cardiovascular death or HF hospitalization, as well as cardiovascular death. Second, from the covariate selection process, we determined that the stress-inducible cytokine, GDF-15, had the strongest association with the primary endpoint and its components. Other markers and characteristics of importance to risk stratification included the biomarkers NT-proBNP, albumin, BUN, and uric acid. Third, whereas there was relative consistency in the prognostic factors affecting the composite outcome and cardiovascular death, those related to HF hospitalization differed substantially: in particular, NT-proBNP did not register among them, underscoring the complexity of both predicting HF hospitalization alone and combining it with cardiovascular death in a primary composite outcome.

The current analytical approach highlights the challenges of pre-selecting risk stratification variables and the important incremental value in the learnings provided by more informed covariate-adjustment in a clinical trial once outcomes have been established but before treatment assignment is revealed. Recent application of the 5-STAR approach by Mehrotra and Marceau West yielded 4 risk strata based on quartiles of NT-proBNP and NYHA class in the VICTORIA trial and also demonstrated a more pronounced treatment effect of vericiguat on cardiovascular death (estimated HR of 0.83, 95% CI 0.71-0.97) than reported in the original primary analysis.(4) The further improvement in the magnitude of that estimate of vericiguat’s effects on cardiovascular death (HR 0.79, 95% CI 0.67-0.93) in the current analysis likely reflects the addition of prognostically relevant biomarkers such as GDF-15. The stronger efficacy signal elucidated with 5-STAR can be attributed to improved risk stratification based on variables observed to be jointly strongly associated with risk for the primary endpoint and its components. By contrast, the original efficacy analyses appeared to have been diluted by pre-selection of stratification factors (region and race) that turned out to be prognostically unimportant.

When applied to the VICTORIA database in the current report, multiple strata were constructed based on the findings that several biomarkers provided the most prognostic information influencing the primary endpoint and both of its components. The novel finding revealing GDF-15 as the single most important variable in the expanded development of the risk strata, accompanied by 4 additional biomarkers, is of interest on several counts. An elevated GDF-15 level reflects activated systematic inflammation, tissue injury, and oxidative stress and is known to be an important determinant of outcomes in high-risk HF patients.(11,12) Several studies have demonstrated that inflammatory markers at baseline correlate with adverse outcomes(7,13–15); however, to our knowledge, this is the first demonstration that GDF-15 constitutes a highly consistent important risk factor in high-risk HFrEF patients.

The finding that NT-proBNP remains a highly important prognostic biomarker for both the composite outcome and cardiovascular death is not unexpected. This biomarker has been shown to be the most important risk marker in the placebo model of VICTORIA as well as in other high-risk HFrEF datasets.(9,16–20) Although NT-proBNP was the second most important variable associated with the primary endpoint and cardiovascular death models, it is noteworthy that it did not appear in the HF hospitalization model. This finding differs from most previous studies and likely reflects the presence and strength of the GDF-15 biomarker and the complexity of predicting the HF hospitalization endpoint alone.(21) It may also indicate the excellent indication-corrected adherence to guideline-based medical therapy that accounted for patient-level indications, contraindications, and tolerance in the trial.(22,23)

Albumin emerged as an important marker of cardiovascular death consistent with other studies and with hypoalbuminemia reflective of abnormal nutritional status, gastrointestinal malabsorption, volume expansion, and systemic inflammation.(22) Uric acid also emerged as an important covariate in the HF hospitalization model and is a known predictor of all-cause death, cardiovascular death, and rehospitalization in patients with HF. Hyperuricaemia has been associated with worse hemodynamic status and contributes to the pathophysiological process of adverse ventricular remodeling, myocardial fibrosis, cardiac hypertrophy, and impaired contractility.(24) BUN also emerged in the HF hospitalization model, a finding that has been shown in multiple other studies,(16,17,25–29) highlighting the importance of the cardio-renal axis in the pathophysiology of HF hospitalization. It has been argued that BUN is not only a reflection of glomerular filtration rate but also an indicator of protein intake, catabolism, and tubular reabsorption.

Using the 5-STAR methodology, the treatment effect of vericiguat was strengthened as indicated by the improved HR for the primary endpoint (from 0.90 to 0.85). Importantly, the treatment effect on cardiovascular death also improved from a non-significant HR of 0.93 (95% CI 0.81-1.06) to a significant HR of 0.79 (95% CI 0.67-0.93) with the risk strata adjustment (**Central Illustration**).

**Central Illustration:**
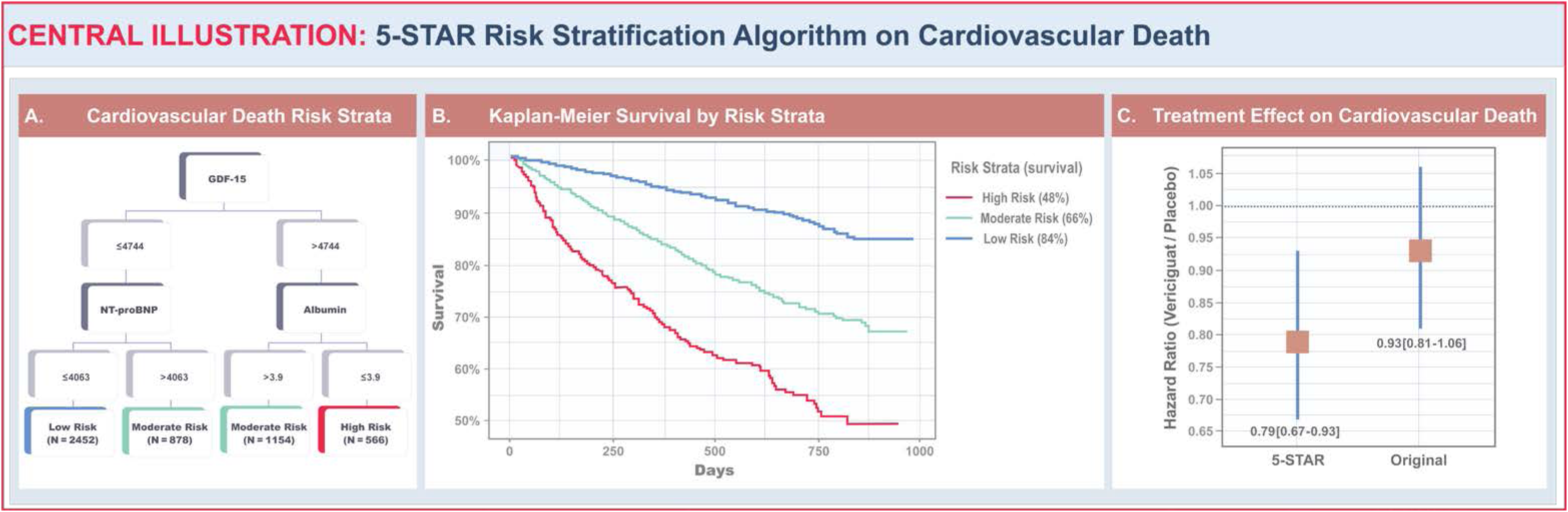
5-STAR Risk Stratification Algorithm for Cardiovascular Death. Panel A: Characteristics defining risk; Panel B: Kaplan-Meier survival estimates for risk strata; Panel C: Vericiguat treatment effect hazard ratio point estimates for cardiovascular death from 5-STAR (left) and original time-to-event analysis (right). 5-STAR, 5-step stratified testing and amalgamation routine; GDF-15, Growth Differentiation Factor 15; NT-proBNP, N-terminal pro-B-type natriuretic peptide.

Finally, the question of vericiguat’s effect in the context of risk heterogeneity is better illuminated in this analysis. Our findings suggest that vericiguat’s benefit may extend across the spectrum of endpoints as demonstrated by the HRs for the composite endpoint and each of its components. This suggests, but does not prove, that the addition of novel baseline biomarker measures provides incremental insights into the role of vericiguat therapy in the high-risk population. Moreover these findings are contrary to the outcomes reported in the prespecified covariate adjustment of the VICTORIA dataset with respect to NT-proBNP quartiles.(2)

Our findings suggest that HF clinical trialists would be wise to collect relevant mechanistic biomarkers and utilize the findings from measurements of inflammation, hemodynamics, and other pathways for risk stratification. Their potentially broader implications may also provide a stimulus for clinical trialists to prospectively evaluate the current methodology in future clinical trials as a complement to standard analysis. In this respect it is noteworthy that the 5-STAR methodology has been prespecified as an ancillary analysis in the ongoing VICTOR trial examining vericiguat in a stable HF population treated with contemporary guideline-based therapy (Ciaran J. McMullan, MBBCh, MMSc, unpublished data, 2024).(30) These findings may also deserve consideration by guideline committees when assessing treatment efficacy.

In the current analysis there is a signal suggesting that the efficacy of vericiguat for the reduction of cardiovascular death may have been attenuated because of dilutional heterogeneity related to the absence of informative covariates in the original analysis. In light of the significant effect on cardiovascular death in the current analysis, it is interesting to speculate that—given the regulatory authorities advocacy of covariate adjustment—had the 5-STAR methodology been prospectively identified as the primary analysis, the recommendations for vericiguat may have been similar to other recently approved HF therapies.(31,32)

### Strengths and Limitations

This study has both strengths and limitations. First our use of the 5-STAR analysis incorporating the mechanistic biomarkers and electrocardiographic data represents the largest and most comprehensive HF database to date utilizing this methodology.. Second, the findings of the novel 5-STAR methodology have demonstrated insights into treatment efficacy of vericiguat in the VICTORIA trial; these hypotheses generating observations are subject to prospective validation. This opportunity may be provided by the forthcoming VICTOR trial and signals the potential utility of integrating a full core laboratory of biomarkers as well as other relevant but as-yet undefined risk covariates into that database.

## Conclusion

Application of the 5-STAR methodology in a large phase 3 clinical trial provides novel insights into the development of treatment-independent risk stratification, thereby attenuating dilution of treatment effects inherent in conventional risk heterogeneity. This retrospective analysis of VICTORIA reveals greater efficacy and more favorable treatment effect for vericiguat versus placebo for the primary endpoint and its two components of cardiovascular death and HF hospitalization. The 5 new covariates associated with outcomes were the biomarkers GDF-15, NT-proBNP, albumin, BUN, and urate, suggesting that inflammatory and hemodynamic pathways are important modulators of vericiguat’s treatment effect. GDF-15 and to a lesser extent NT proBNP were the most important risk markers and their relative roles demonstrate the complexity of the relationships between cardiovascular death and heart failure-related hospitalization which are commonly combined within a composite endpoint. Confirmation of these findings may stimulate revisiting recommendations for the treatment of patients with HFrEF with vericiguat. We contend this methodology deserves consideration in future clinical trials and by guideline committees and regulatory agencies evaluating novel therapies.

## Data Availability

Data Availability Statement
Requests to access participant-level data from qualified researchers trained in human subject confidentiality protocols and in accordance with the VICTORIA data-sharing charter may be submitted at https://thecvc.ca/victoria/data-sharing/.

## PERSPECTIVES

### Competency in Medical Knowledge

The 5- STAR methodology attenuates the dilution of effects inherent in prognostic risk heterogeneity characterizing conventional clinical trials.

### Competency in Patient Care

Novel biomarkers characterizing inflammatory and hemodynamic pathways and GDF-15 in particular, enhance the understanding of treatment - independent risks of cardiovascular death and the composite outcome of CV death and heart failure hospitalization in patients with HFrEF thereby adding new insights into the potential efficacy of vericiguat.

### Translational Outlook Implication

Using the 5-STAR methodology in concert with the incorporation of novel biomarkers deserves consideration in future clinical trials and by guideline committees and regulatory agencies evaluating novel therapies.

## Acknowledgments

We are pleased to recognize Wendimagegn Alemayehu, PhD for his assistance in the statistical analysis and critical review of the manuscript.

Elizabeth E.S. Cook of the Duke Clinical Research Institute and Lisa Soulard of the Canadian VIGOUR Centre provided editorial support.

## Non-standard Abbreviations and Acronyms

BUN: blood urea nitrogen
CI: confidence interval
GDF-15: growth differentiation factor-15
HF: heart failure
HFrEF: heart failure with reduced ejection fraction
HR: hazard ratio
ICD: implantable cardioverter defibrillator
NYHA class: New York Heart Association Heart Failure Classification System
NT-proBNP: N-terminal pro-B-type natriuretic peptide
5-STAR: 5-step stratified testing and amalgamation routine

