## Supplemental Materials for "Assessing Benefit in Heart Failure Patients with Reduced Ejection Fraction: Analysis of the VICTORIA Trial Using Novel Prognostic Risk Stratification"

### SUPPLEMENTAL MATERIAL

**Table S1.** Candidate Variables for the 5-STAR

| Model | Category | Variables included |
|---|---|---|
| Mentz (66 variables) | Trial details | Subject randomized while hospitalized |
| | Demographic details | Age, sex, region/race (incorporation of race for NA)*, ethnicity |
| | HF details | Index event, time from index event to randomization, duration of HF diagnosis, duration of primary diagnosis of HFrEF, ejection fraction, NYHA class |
| | Past medical history | CCSA class, atrial fibrillation, atrial flutter, COPD, angina, diabetes, hypertension, hyperlipidemia, anemia, sleep apnea, peripheral artery disease, prior MI, prior stroke, prior TIA, prior CABG, prior PCI, aortic valve replacement, mitral valve replacement, current tobacco use |
| | Physical exam/vitals | BMI, height, weight, systolic blood pressure, diastolic blood pressure, pulse pressure |
| | Medications and devices | ACE or ARB, beta blocker, MRA, sacubitril-valsartan, ivabradine, ICD, pacemaker |
| | Laboratories | Creatinine, eGFR, NT-proBNP, hemoglobin, potassium, sodium, albumin, ALT, AST, bicarbonate, bilirubin, BUN, calcium, chloride, GGT, glucose, hematocrit, platelets, white blood count, red blood count, urate |
| | ECG | QTcF |

| | | |
|---|---|---|
| deFilippi (5 variables) | Biomarkers | hsCRP, Cystatin C, GDF-15, IL-6, hs-TNT |
| Yogasundaram (11 variables) | Electrocardiographic measures | sinus rhythm, afib/flutter, tachycardia, heart rate, QRS duration, RBBB, LBBB (conventional), LBBB (Strauss), LBBB (conv/Strauss), paced rhythm, QTc/ms |

*Race/Region included as separate variables

5-STAR, 5-step stratified testing and amalgamation routine; NA, North America; HF, heart failure; HFrEF, HF with reduced ejection fraction; NYHA, New York Heart Association; CCSA, Canadian Cardiovascular Society angina; COPD, chronic obstructive pulmonary disease; MI, myocardial infarction; CABG, coronary artery bypass graft; TIA, transient ischemic attack; PCI, percutaneous coronary intervention; BMI, body mass index; ACE, angiotensin-converting-enzyme; ARB, angiotensin ii receptor blocker; MRA, mineralocorticoid receptor antagonists; ICD, implantable cardioverter-defibrillator; eGFR, estimated glomerular filtration rate; NT-proBNP, N-terminal pro-B-type natriuretic peptide; ALT, alanine transaminase; AST, aspartate aminotransferase; BUN, blood urea nitrogen; GGT, gamma-glutamyl transferase; ECG, electrocardiogram; QTcF, QTc corrected using Fridericia's formula; hsCRP, high-sensitivity C-reactive protein; GDF-15, Growth Differentiation Factor 15; IL-6, interleukin 6; hs-TNT, hs-TNT, high-sensitivity Troponin T; afib, atrial fibrillation; RBBB, right bundle branch block; LBBB, left bundle branch block

**Figure S1.** Tree with Final Risk Strata Assignments for the Composite Endpoint of Cardiovascular Death or HF Hospitalization. HF, heart failure; GDF-15, Growth Differentiation Factor 15; NT-proBNP, N-terminal pro-B-type natriuretic peptide.

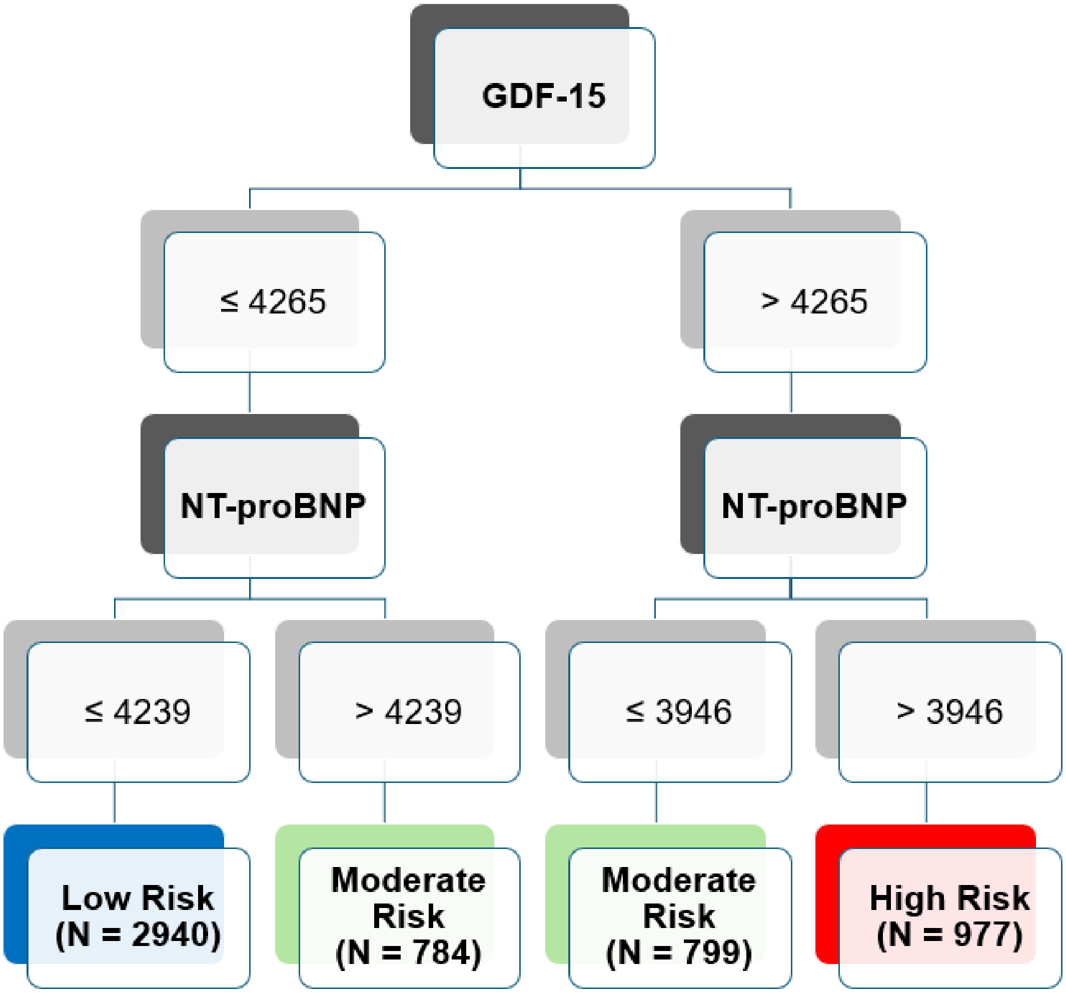

**Figure S2.** Tree with Final Risk Strata Assignments for the Endpoint of Cardiovascular Death. GDF-15, Growth Differentiation Factor 15; NT-proBNP, N-terminal pro-B-type natriuretic peptide.

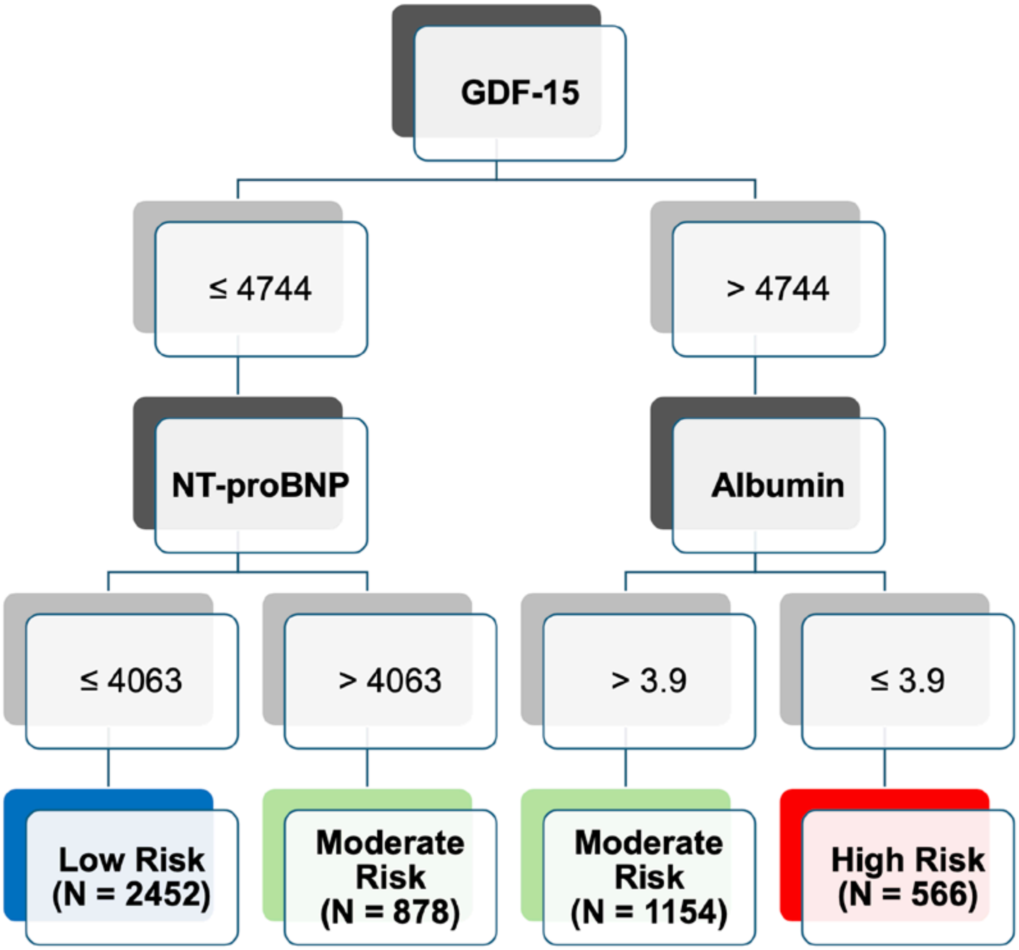

**Figure S3.** Tree with Final Risk Strata Assignments for Endpoint of HF Hospitalization.

HF, heart failure; GDF-15, Growth Differentiation Factor 15; BUN, blood urea nitrogen.

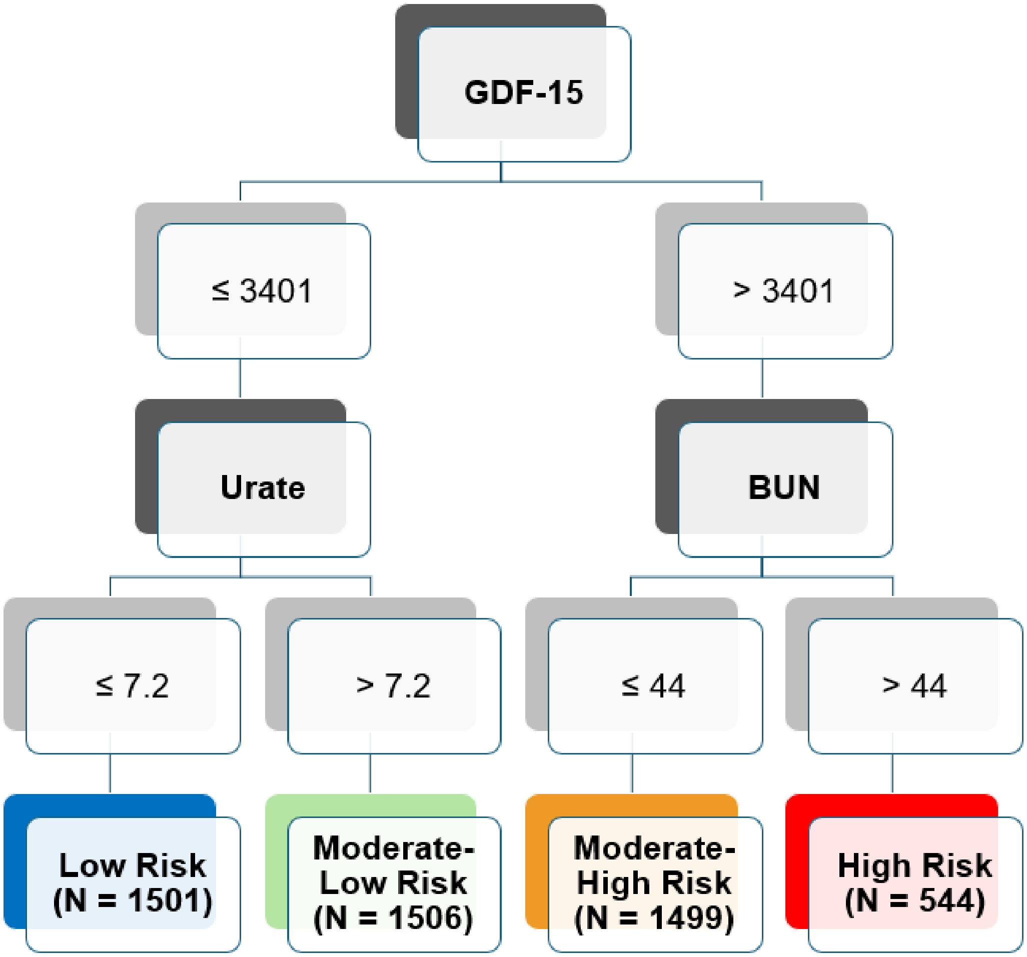
